# Prediction of Covid-19 Infections Through December 2020 for 10 US States Incorporating Outdoor Temperature and School Re-Opening Effects-October Update

**DOI:** 10.1101/2020.11.14.20231902

**Authors:** Ty Newell

## Abstract

Control of SARS-CoV-2 transmission requires control of two human behaviors. A two-parameter, human behavior Covid-19 infection growth model continues to accurately predict total infections based on gross human interaction and local human interaction behaviors for 10 US States (NY, WA, GA, IL, MN, FL, OH, MI, CA, NC). Since prediction model initiation on July 27, 2020, total infections for 8 states have grown by more than 200%. Only New York (23% infection growth) and Florida (189% infection growth) have grown less.

October displays combined impacts of increased social interactions as schools and businesses increase physical gatherings, coupled with climate dependent local interactions. The US, on average as of the end of October, has an Infection Parameter (IP) of 3.4 representing accelerating infection growth. Gross human interactions must be reduced by 15% or local interaction behavior (eg, face mask usage, ventilation) must be improved to reduce disease transmission efficiency by 27% in order to reach the linear infection growth boundary separating accelerating infection growth from decaying infection growth regions.

Eastern States (NY and NC) have had mild fall temperatures, which increases outdoor activities and increases building fresh air ventilation rates that suppress virus transmission efficiency. Mild temperatures in southern States (GA, FL and CA) during October have also helped suppress virus transmission. Midwest States experienced highly elevated infection rates due to combined effects of school openings coupled with a truncated fall season. WA stayed in the beneficial 70F (22C) to 50F (10C) zone through October, with minimal accelerated infection growth, but is now entering its heating season with average outdoor temperatures below 50F that are contributing to increased disease transmission efficiency.

## Introduction

SARS-CoV-2 disease transmission modeling predictions as of October 31, 2020 are reported for 10 US States (NY, WA, GA, IL, MN, FL, OH, MI, CA, NC) since model initiation on July 27, 2020, extending through December 31, 2020. The ten US States selected for prediction modeling display a wide variety of infection growth characteristics. Despite the variation of infection growth patterns and infection levels that differ by an order of magnitude, the two-parameter, behavior model accurately predicts actual infection numbers for all ten US States.

Control of SARS-CoV-2 transmission is a function of two independent human behavior characteristics. Gross human interaction, defined by a Social Distance Index (SDI) parameter, is a measure of the extent of one’s travel outside of their home, such as traveling to school, work, or shopping. Local human interaction, defined by a virus transmission efficiency (G) describes interpersonal interactions among people in a shared space such as “distancing”, face mask usage, hugging, fresh air ventilation, air filtration, and other factors that impact how SARS-CoV-2 is transmitted among people. Outdoor temperature impacts local disease transmission efficiency because inclement conditions encourage more indoor activities.

Four prediction cases for each State represent combinations of the two independent human behaviors.

1. no school with outdoor temperature effect
2. no school with no outdoor temperature effect
3. school with temperature effect
4. school with no temperature effect

Throughout most of the US, infections grew at an accelerated pace during October. Infection growth is due to a combination of increased social interactions such as physical school openings and increased disease transmission efficiency caused by increased indoor gatherings from cold weather coupled with relaxed distancing, decreased face mask usage, and other “Covid fatigue” behaviors.

Reduction of gross social interactions by 15% or reduction of local human behavior practices by 27% are required to return to the linear infection growth boundary separating accelerating and decaying infection growth regions. Sustained reduction of both behaviors below the linear infection growth boundary are necessary for decreasing infections.

## Background

Two human behaviors define an Infection Parameter (IP) that is similar to the basic Reproduction number, Ro. IP relates infection growth to current number of infections and is conceptually the number of new infections over a two week period per infectious person. An IP value of 2.72 (“e”) results in linear infection growth. Linear infection growth is a boundary separating accelerating and decaying infections. The prediction model has been described in detail (1,2) with monthly prediction updates since the July 27 model initiation for 10 US States (3,4).

A key assumption in infection prediction is an observation of human tendency to gravitate to the linear infection growth boundary separating increasing and decreasing new daily infections. In the absence of a coherent, sustained virus control plan, a populace tends to alternately move between accelerating infection growth and decaying infections. The reason for the push-pull across the linear growth boundary is assumed to be caused by relaxed human behavior when infections are receding that increases IP above 2.72, followed by a populace reacting to reports of uncontrolled infection growth.

Figure 1 is a schematic showing how gross and local human behaviors affect the Infection Parameter, IP. The “No Temp” and “No School” statepoint in Figure 1 represents an initial condition of many States as the summer surge was suppressed and various regions returned to linear infection growth with an IP of 2.72 at the end of July when the model was initiated. The four case conditions modeled for each State are shown in Figure 1. “School” represents increased movement outside of the home, which is a “gross” human interaction behavior. Associated with school openings would be increased work activities by adults as children return to school and daycare facilities. “Temperature” represents movement into or out of building environments, and is a local human behavior that impacts disease transmission efficiency. Between 50F (10C) and 70F (22C) people are more active outside, which reduces disease transmission. Buildings are better ventilated within this outdoor temperature range, too, and more outdoor activities results in less contagion concentration indoors.

**Figure 1.**
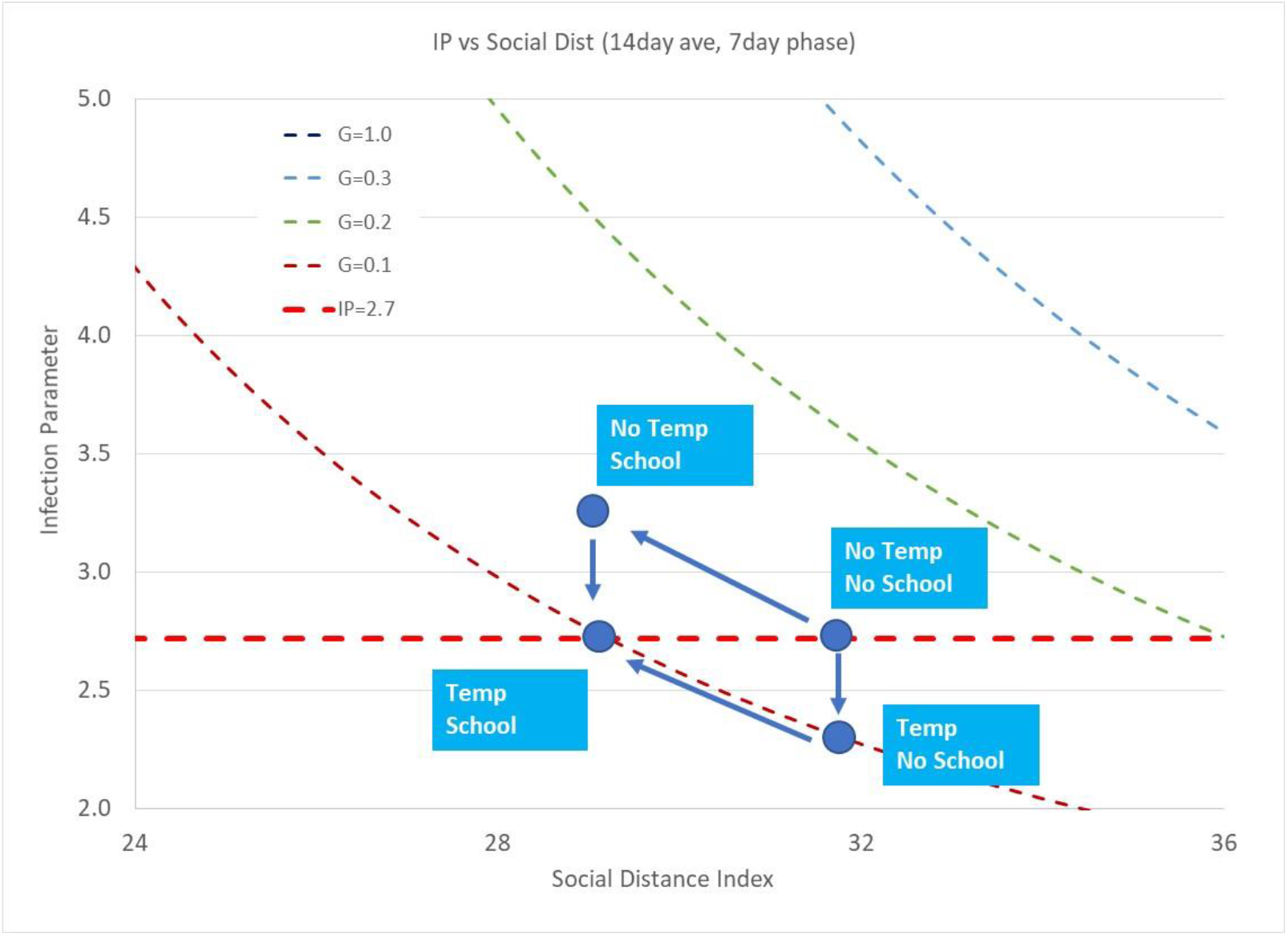
Schematic showing example IP paths for school opening and fall temperature effects on gross human behavior (Social Distance Index) and virus transmission efficiency (G).

Movement in the IP-SDI-G space is path dependent. Figure 1 shows two example paths that begin on the linear growth IP boundary with no school opening or outdoor temperature impacts. More northerly regions could move from warm summer conditions (outdoor temperature above 70F (22C)) to the beneficial swing season conditions between 50F (10C) and 70F (22C) prior to school openings and increased business/social interactions. Local disease transmission efficiency (G) decreases without a change in the Social Distance Index (SDI), decreasing IP below the 2.72 linear growth boundary, resulting in decreasing new daily infections.

In contrast to IP movement due to beneficial outside temperature changes that affect local interaction behavior, gross human interaction variation is a parallel movement along an iso-disease transmission efficiency curve. Increased movement of people outside their homes is a decrease in SDI, a parameter obtained from the Maryland Transportation Institute and the University of Maryland (5) based on anonymous cell and vehicle gps data. Reduction of SDI increases IP above the linear infection growth boundary, resulting in accelerating infection growth.

Actual movement in IP is a complex interaction of gross and local behaviors, with resulting infections dependent on the path taken from one statepoint to another. Following the example points in Figure 1, a location that first experiences a drop in IP due to beneficial outdoor temperatures, followed by increased social movement (decreased SDI) will accumulate a different amount of infections in comparison to a location that first experiences a reduction in SDI followed by a beneficial change outdoor temperature or other beneficial local human behavior (eg, increased face mask usage). The Infection Parameter end point as a result of both SDI (gross human behavior) and G (local human behavior) movements may be the same for each path, but resulting infections are not.

Detailed explanation of the human behavior prediction model is included in reports (1) and (2), with update reports describing trends for August (3) and September (4). All data used for model development and model comparison are available to the general public (5, 6, 7).

### Comparison of End-of-October Actual Infections and Predicted Infections

Table 1 lists actual Covid-19 total infections and predicted total infections as of October 31, 2020. The prediction model was initialized as of July 27, 2020 (2). Four case predictions for each State have been made:

**Table 1.**
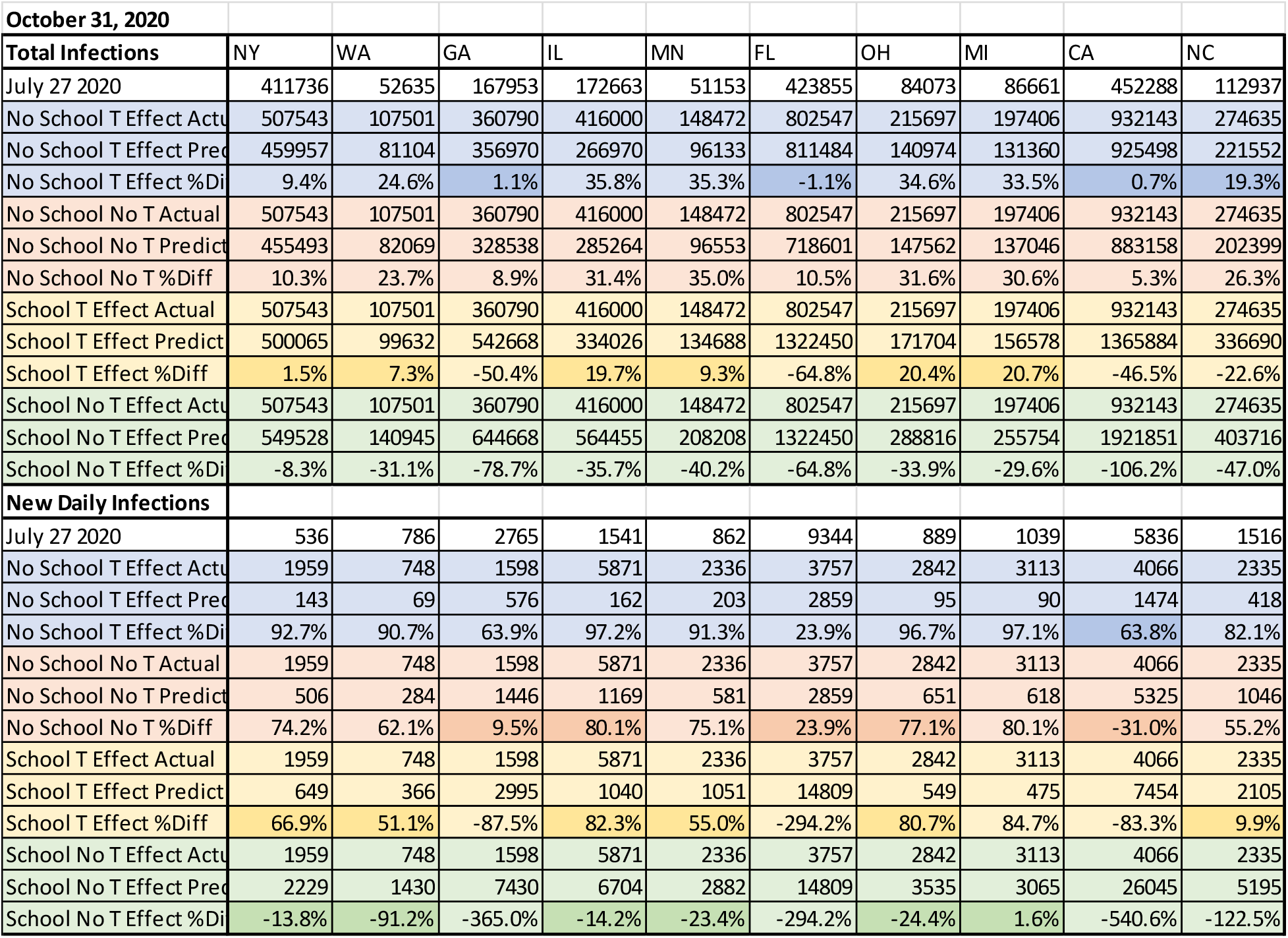
Total Infections and New Daily Infections as of July 27, 2020 October 31, 2020 actual and October 31, 2020 predictions for 10 US States.

1. no school with outdoor temperature effect
2. no school with no outdoor temperature effect
3. school with temperature effect
4. school with no temperature effect

Total infection predictions for all 10 States fall within the boundaries of the four case conditions modeled, similar to the IP region shown in Figure 1. That is, some case results overestimate predicted infections (negative percentages) while other predictions underestimate infections (positive percentages). Actual infections fall within a region surrounded by the assumed cases.

Daily new infections are also within the boundaries of the assumed cases, however, the variation between actual new daily cases and predicted are larger than total infection comparisons. The difference between total infections from one day to the next (ie, new infections) is a small number compared to total infections, while daily new infection reports have large day-to-day variations.

### October Outdoor Temperature Effects

Figures 2, 3 and 4 show outdoor temperatures in comparison to the prediction model’s assumed outdoor temperature based on long term average temperature for each region. Ambient temperature assumptions are described in more detail in previous reports (2,3,4).

**Figure 2.**
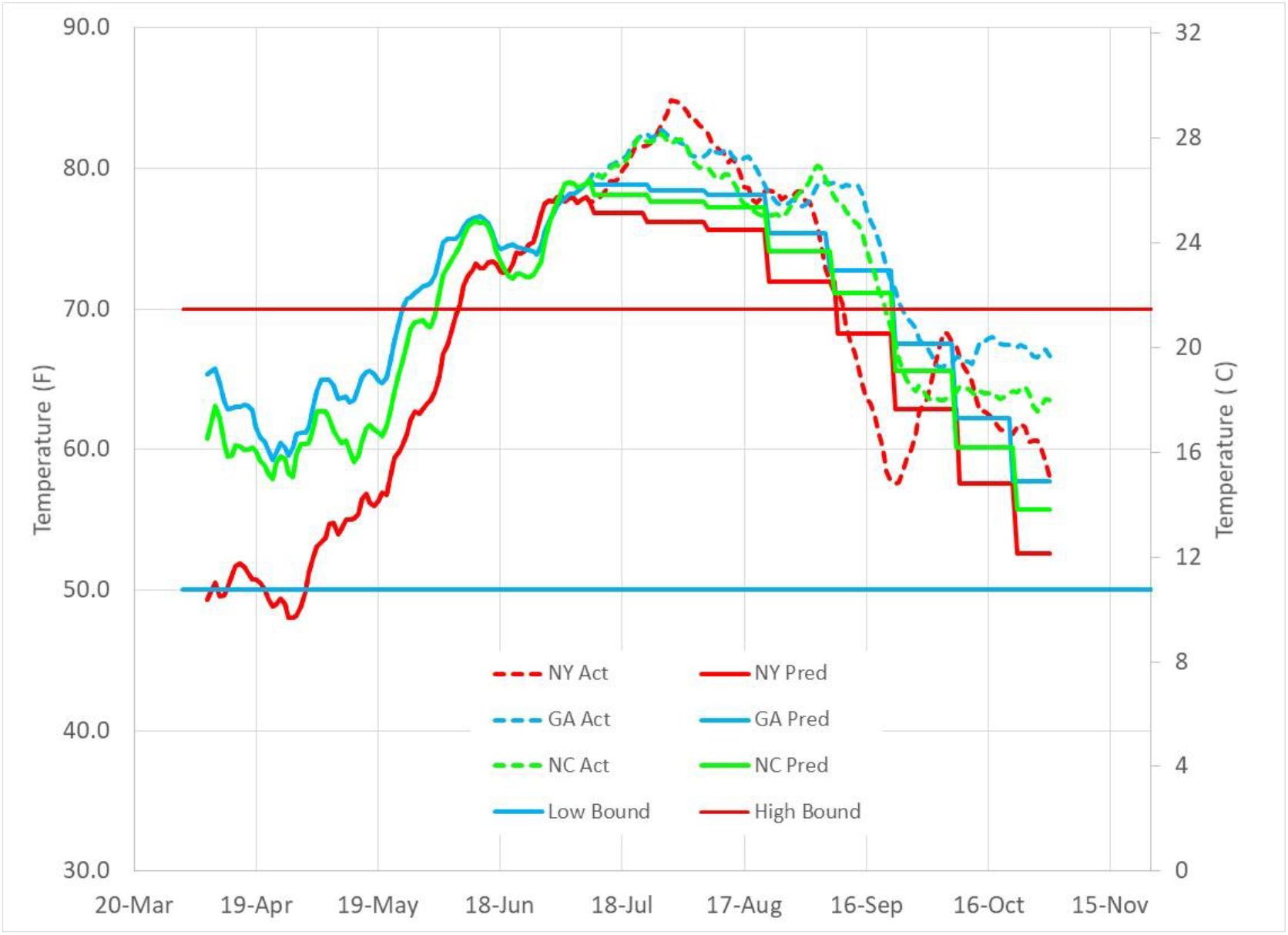
Historical average outdoor temperatures and actual average outdoor temperatures for NY, NC and GA.

**Figure 3.**
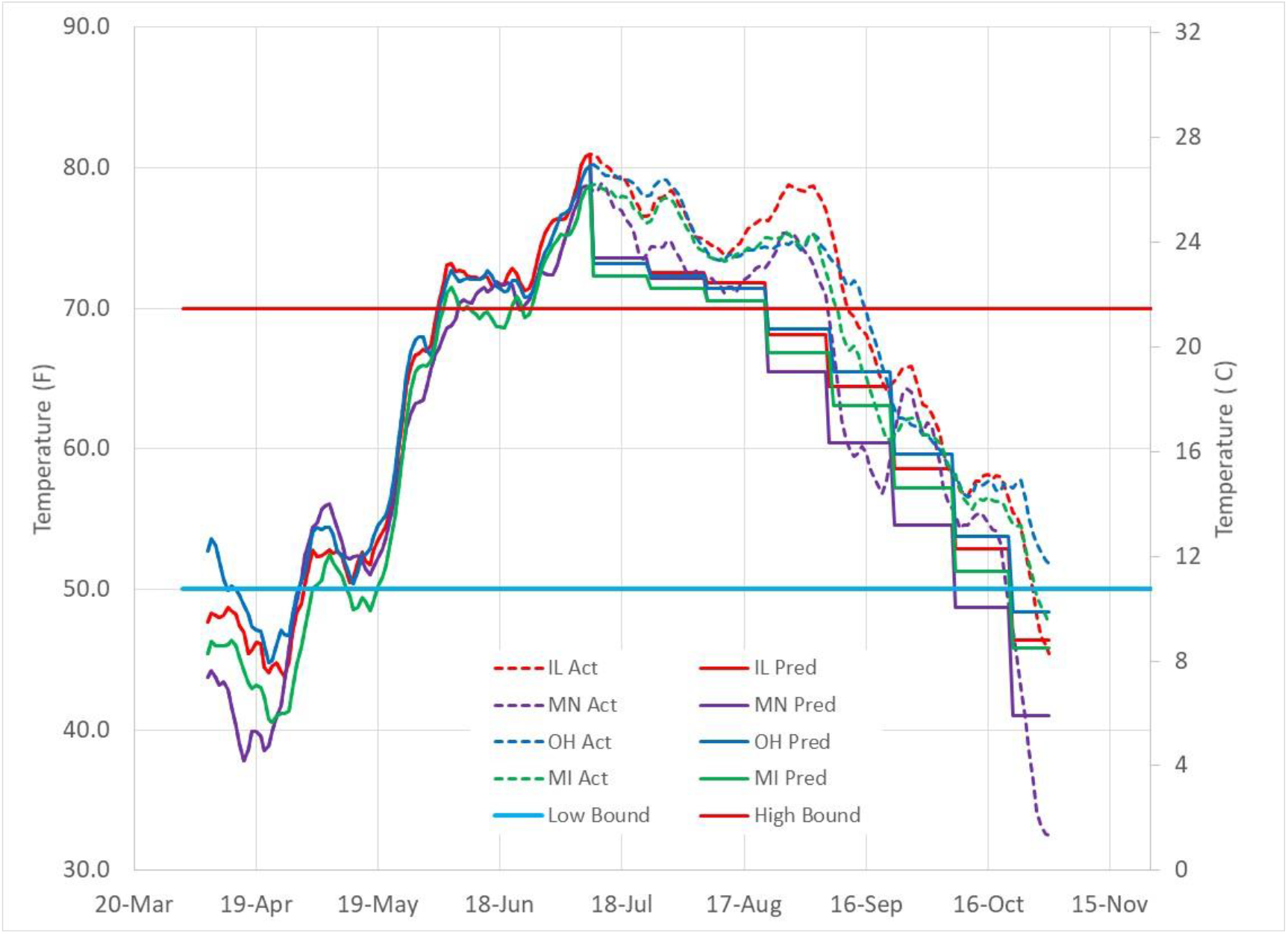
Historical average outdoor temperatures and actual average outdoor temperatures for IL, MN, OH, and MI.

**Figure 4.**
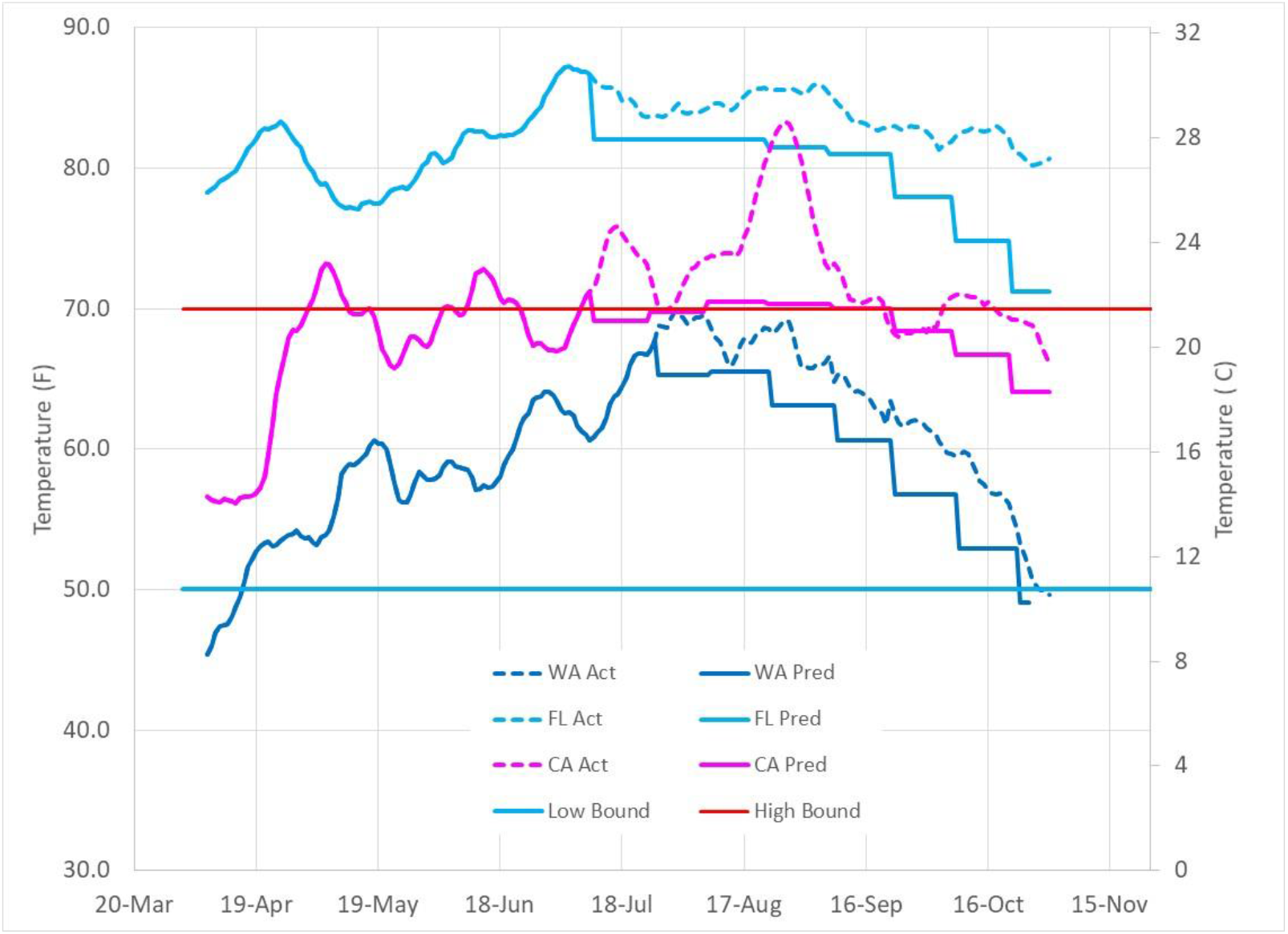
Historical average outdoor temperatures and actual average outdoor temperatures for FL, CA, and WA.

Figure 2 shows that eastern States entered the beneficial swing season temperature range similar to the model’s assumed temperatures. Within the beneficial temperature region, disease transmission efficiency, G, is assumed to decrease by 30% relative to outdoor temperatures that are greater than 70F (22C) or less than 50F (10C). During October, NY, NC and GA have stayed in the beneficial temperature band, helping to reduce infection transmission as people are able to enjoy outdoor environments as well as increase fresh air ventilation in buildings and homes. Note that more time spent outdoors reduces contagion concentration in buildings, too.

Figure 3 shows outdoor temperature trends for Midwestern States (IL, MI, MN, OH). Extended warm conditions resulted in a 3 week delay for entering the beneficial temperature region. Temperatures dropped below 50F (10C) at near normal dates. The truncated swing season reduced the amount of beneficial infection suppression expected for Midwestern States.

Figure 4 shows outdoor temperatures for States that do not tend to move across the beneficial outdoor temperature region. Washington, for example, does not tend to move into elevated (above 70F (22C)) summer temperatures requiring air conditioning. Instead, much of WA enjoys cool summers with enhanced building ventilation and outdoor activities. California (represented by Los Angeles weather data) tends to move along the upper temperature boundary of the beneficial temperature region. Florida, in contrast to WA, tends to stay above the upper 70F (22C) temperature limit for much of the fall, and mostly stays above the lower temperature boundary (50F (10C)) during the winter.

Note that the temperature boundaries and building characteristics are quite variable for different regions. Late spring and early summer 2020 infection trends show a strong linkage to outdoor temperature (1). During this time, social interactions and interaction variations were reduced as much of the US was in “lockdown”, allowing temperature effects to be identified more easily. Fall infection trends are more complex because of varying social interactions as schools and businesses increase physical interactions coupled with temperatures moving into and out of swing season temperatures.

### Infection Parameter (IP) Trends

Figure 5 shows updated Infection Parameter (IP) trends through October 31, 2020 for the 10 modeled States. An important observation is a general tendency for a region to gravitate to the linear infection growth boundary defined by an IP of 2.72 when there are no sustained infection control guidelines. A populace oscillates across the linear growth boundary as it reacts to news of accelerated, out-of-control infection growth by self-enacting infection control (eg, isolation, face mask usage). As IP is reduced below 2.72, new infections decrease, resulting in relaxation of infection controls by a populace until the linear infection growth boundary is crossed again.

**Figure 5.**
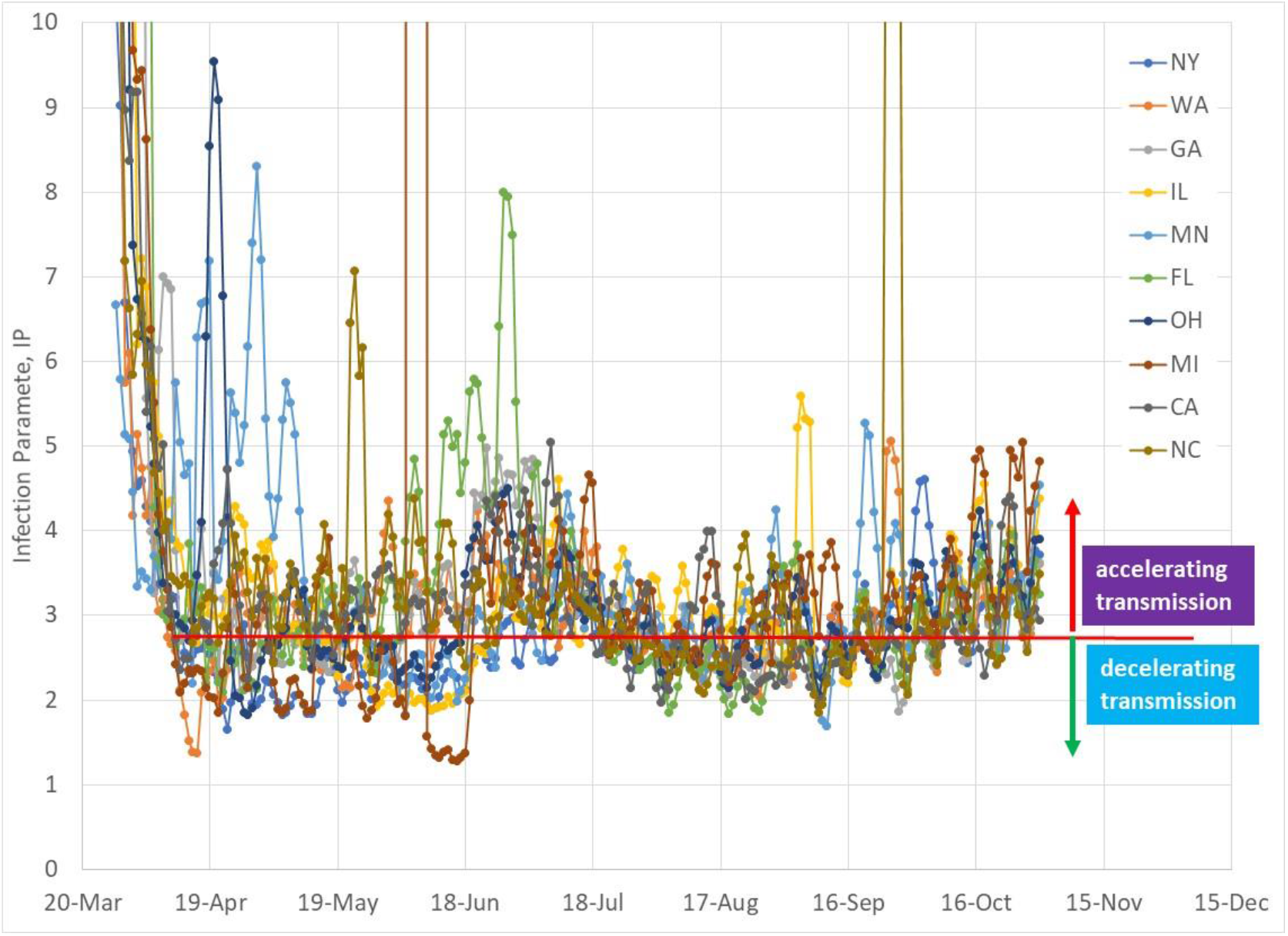
Infection Parameter for 10 US States through October 31, 2020.

As seen in Figure 5, April through May, 2020 were relatively stable periods of linear infection growth. Mid-June and July moved into accelerated infection growth due to a combination of northern States outdoor temperature elevating above 70F (22C) and southern States unwisely re-opening businesses and public gatherings. Late July through mid-September, 2020 are also characterized by a return to linear infection growth as the summer surge was contained with increased disease transmission control measures. By mid-September, temperatures decreasing below 50F (10C) causing increased indoor occupation coupled with physical school openings started increasing IP levels above the linear growth boundary again.

Note that an increasing IP trend could be observed in early September, and that one could project its path several weeks before October’s accelerated infection growth. IP has Covid-19’s incubation period (assumed to be one week) and infectious period (assumed to be two weeks) built into its definition (1,2).

As of the end of October, the average US IP is 3.4, indicating accelerating infection growth with an SDI of 29 and a disease transmission efficiency, G, of 0.15. Returning to the linear infection growth boundary separating accelerating and decaying infection growth regions requires a 15% increase of SDI to 33 or a 27% decrease of the disease transmission efficiency, G, to 0.11. Decreasing IP below 2.72 is required for infection decay, with decay rates increasing with lower IP values. An IP of 1.0 is the lower limit in which either perfect isolation (SDI = 100, no gross human interaction) or zero disease transmission efficiency (G=0, no transmission among people when interacting, eg, a perfect face mask that captures all contagion of emitters and prevents ingestion of contagion by susceptible) results in the fastest infection decay rate (approximately 3 weeks for new daily infections to reach zero).

### October State Infection Growth Trends

Figures 6 through 15 show total infection growth trends for each of the 10 States modeled from July 27 through December 31, 2020. Actual infection data for August, September, and October have been added to the plots. The plots show the four prediction cases:

**Figure 6.**
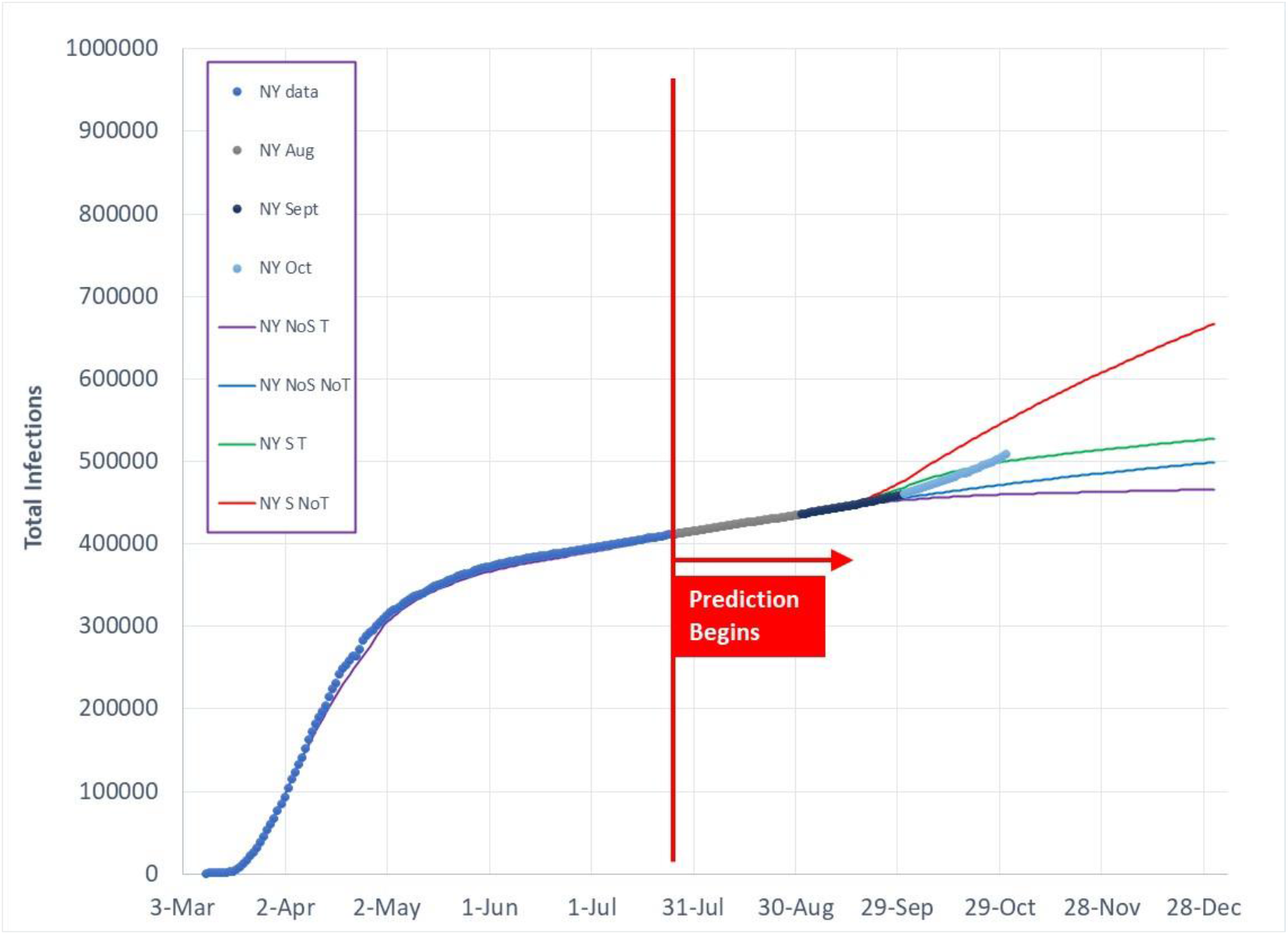
Total Covid-19 infection predictions as of July 27, 2020 for 4 model cases in comparison to actual data for NY.

1. no school with outdoor temperature effect
2. no school with no outdoor temperature effect
3. school with temperature effect
4. school with no temperature effect

October is a month in which the four case paths diverge as climate differences and physical interactions from school and other activities increased. The prediction model assumes increased human interaction during September reduced the State’s Social Distance Index (SDI) by 20%. As discussed in the original infection prediction report (2), a 20% SDI reduction results in a decreased SDI that is somewhat greater than pre-Covid SDI levels. SDI is returned to August SDI levels beyond September to represent populace reaction to accelerated infection growth. The dynamic response of a populace has not been analyzed and is a subject for future research to more properly model transient effects of human behavior.

Note that school openings, and other increased social interactions such as return to work, are not strictly defined by September as many regions within various States phased transitioned gradually from virtual to physical school openings throughout September and October. Combined effects of cold temperatures (below 50F/10C) and increased social interactions resulted in accelerating infection growth are causing schools to virtual classrooms as of the end of October and early November.

Figures 6, 7, and 8 show trends for NY, GA, and NC, respectively. NY (Figure 6) is moving into the infection growth region that reflects decreasing outdoor temperature with increased social interactions. GA (Figure 7) is characterized by two primary paths during October because its weather stays above 70F (22C) through much of October. The two primary paths represent increased social interaction (ie, school openings) and no change in social interactions from those observed in September. As of the end of October, GA appears to have moved through the month with caution as it has largely followed the “no school” prediction paths (ie, no significant reduction of SDI). NC (Figure 8) delayed physical school openings until October 5, and has left physical school openings to local school districts. NC infection path is between the paths followed by NY and GA, showing a gradual trend during October from “no school” cases to “school” (reduced SDI).

**Figure 7.**
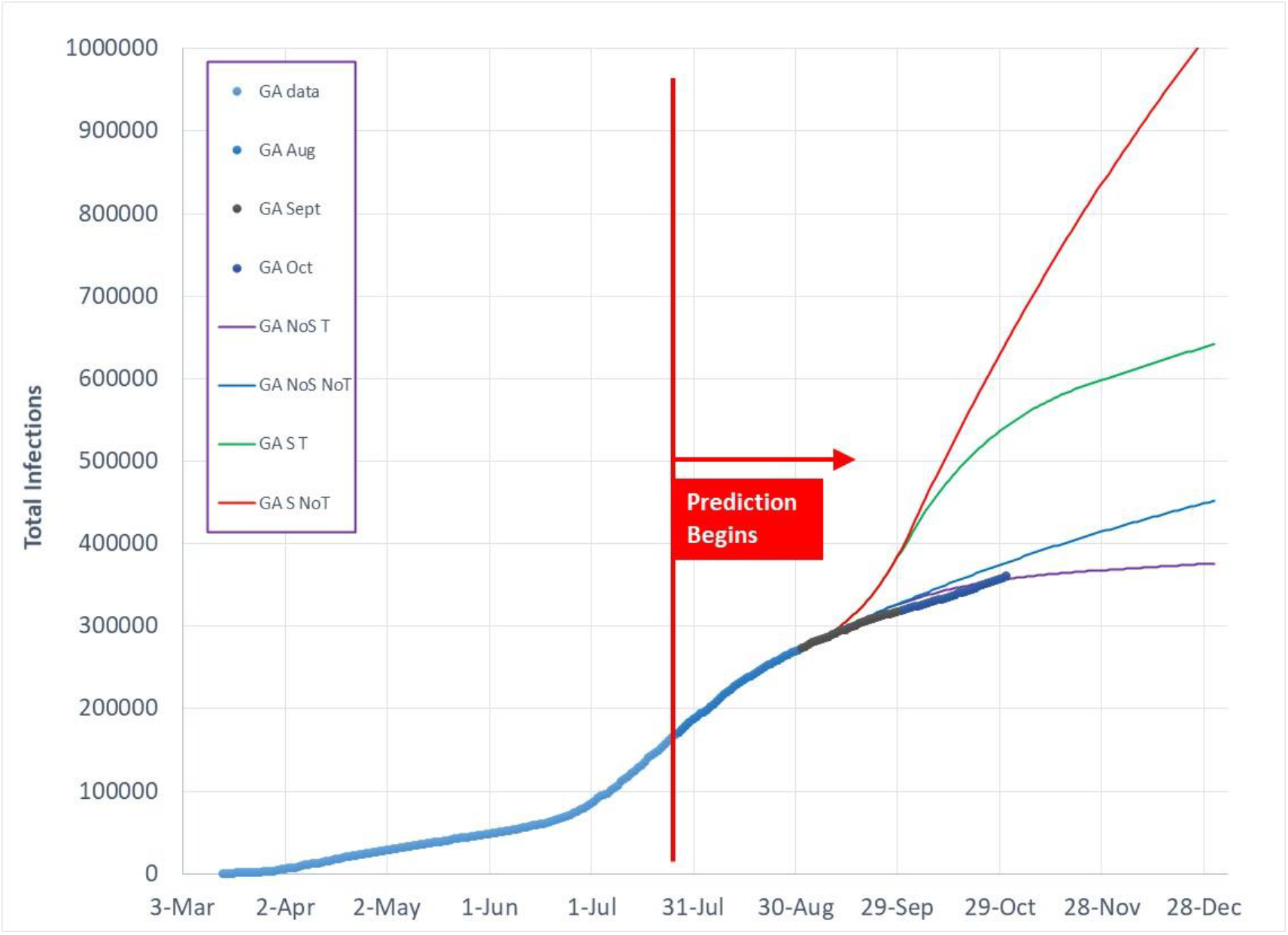
Total Covid-19 infection predictions as of July 27, 2020 for 4 model cases in comparison to actual data for GA.

**Figure 8.**
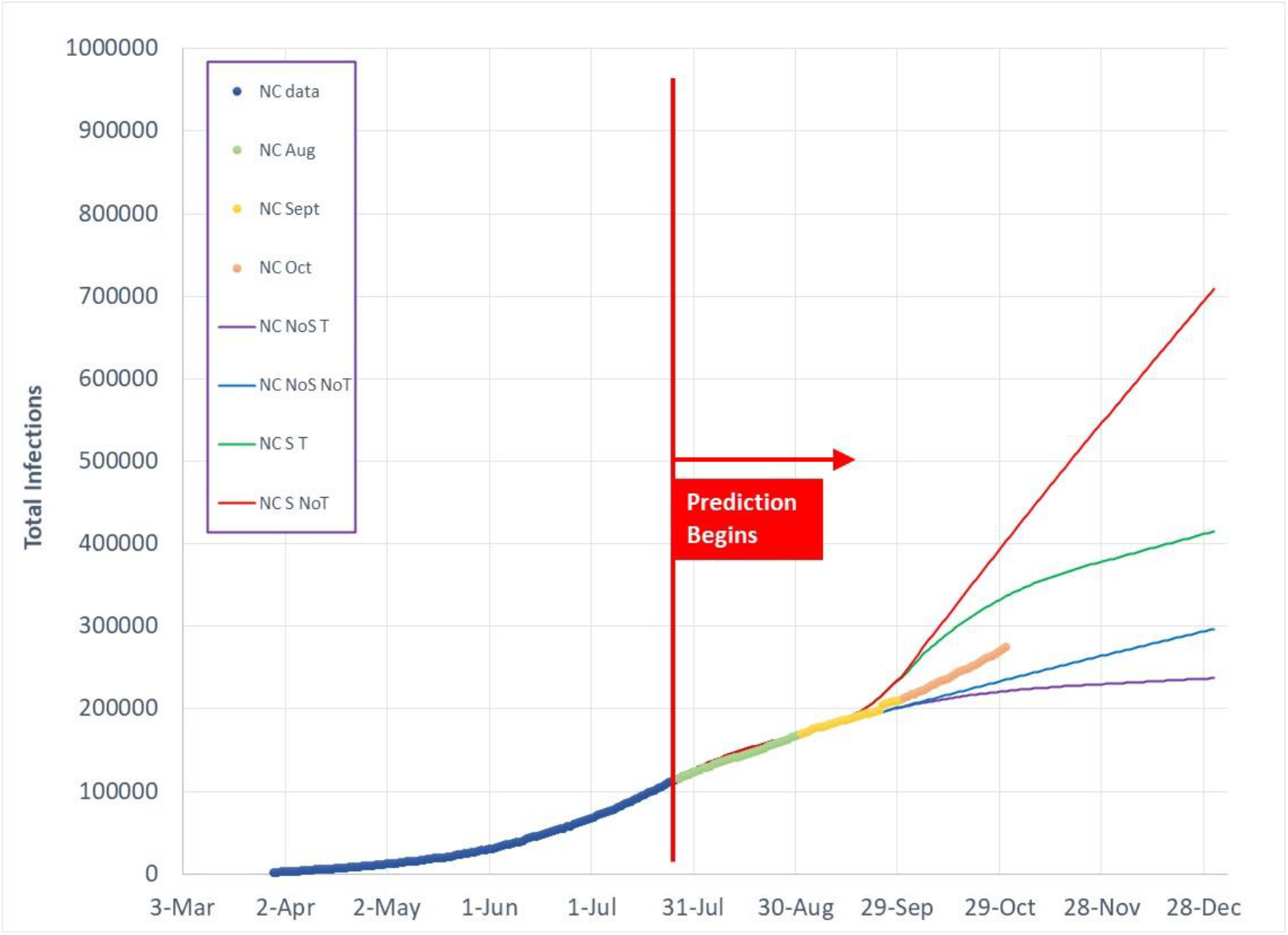
Total Covid-19 infection predictions as of July 27, 2020 for 4 model cases in comparison to actual data for NC.

Figures 9, 10, 11, and 12 show total infection trends for IL, MN, MI, and OH, respectively. All four Midwestern States had shorter swing season temperatures with reduced beneficial suppression of infection transmission, resulting in an infection growth somewhat between school opening cases with and without outdoor temperature effects. Schools and businesses are returning to virtual activities as of the beginning of November. If reactions to accelerating infection growth are insufficient, November’s infection cases may exceed the upper infection case (School opening without beneficial temperature effect) predictions.

**Figure 9.**
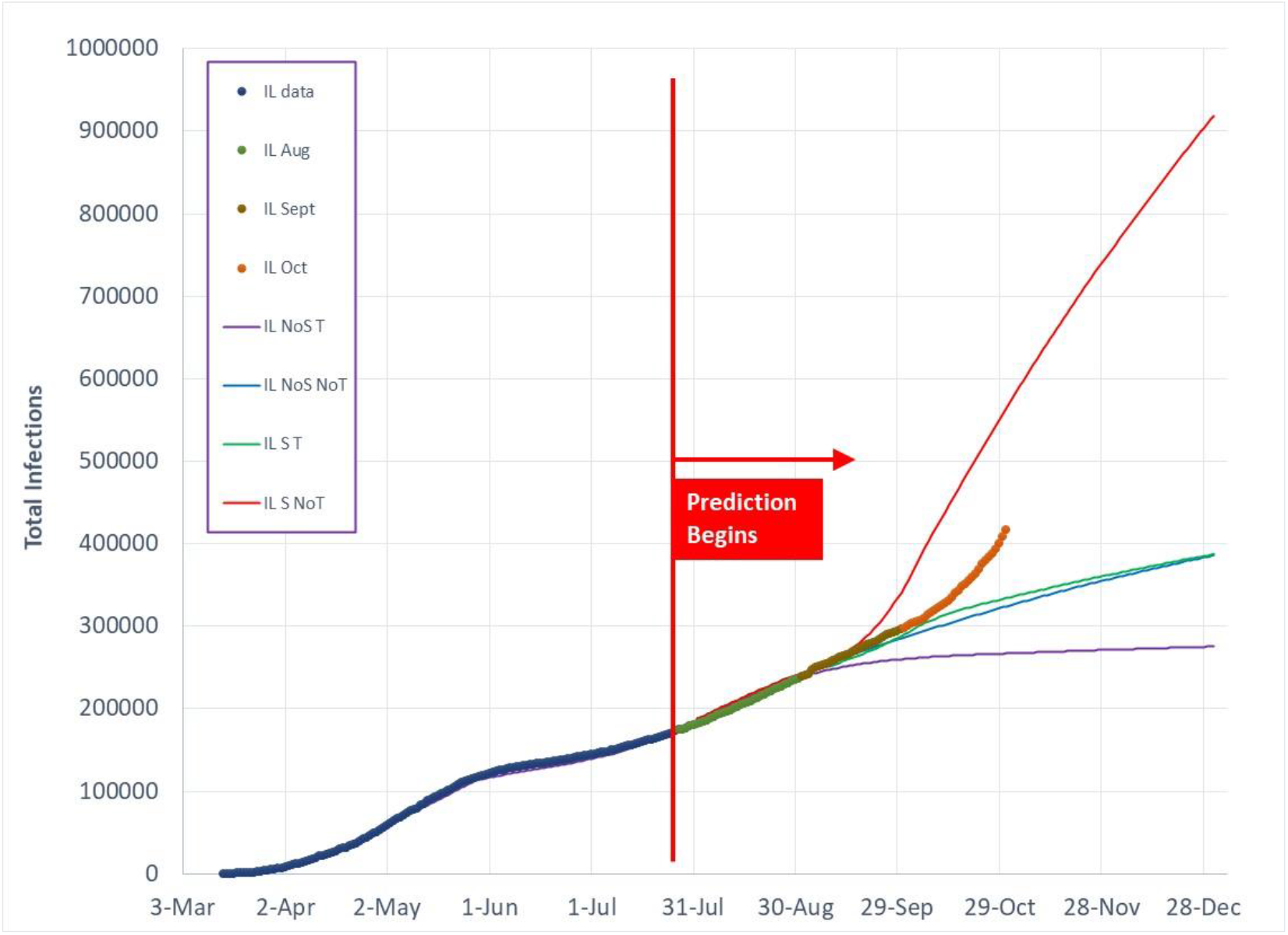
Total Covid-19 infection predictions as of July 27, 2020 for 4 model cases in comparison to actual data for IL.

**Figure 10.**
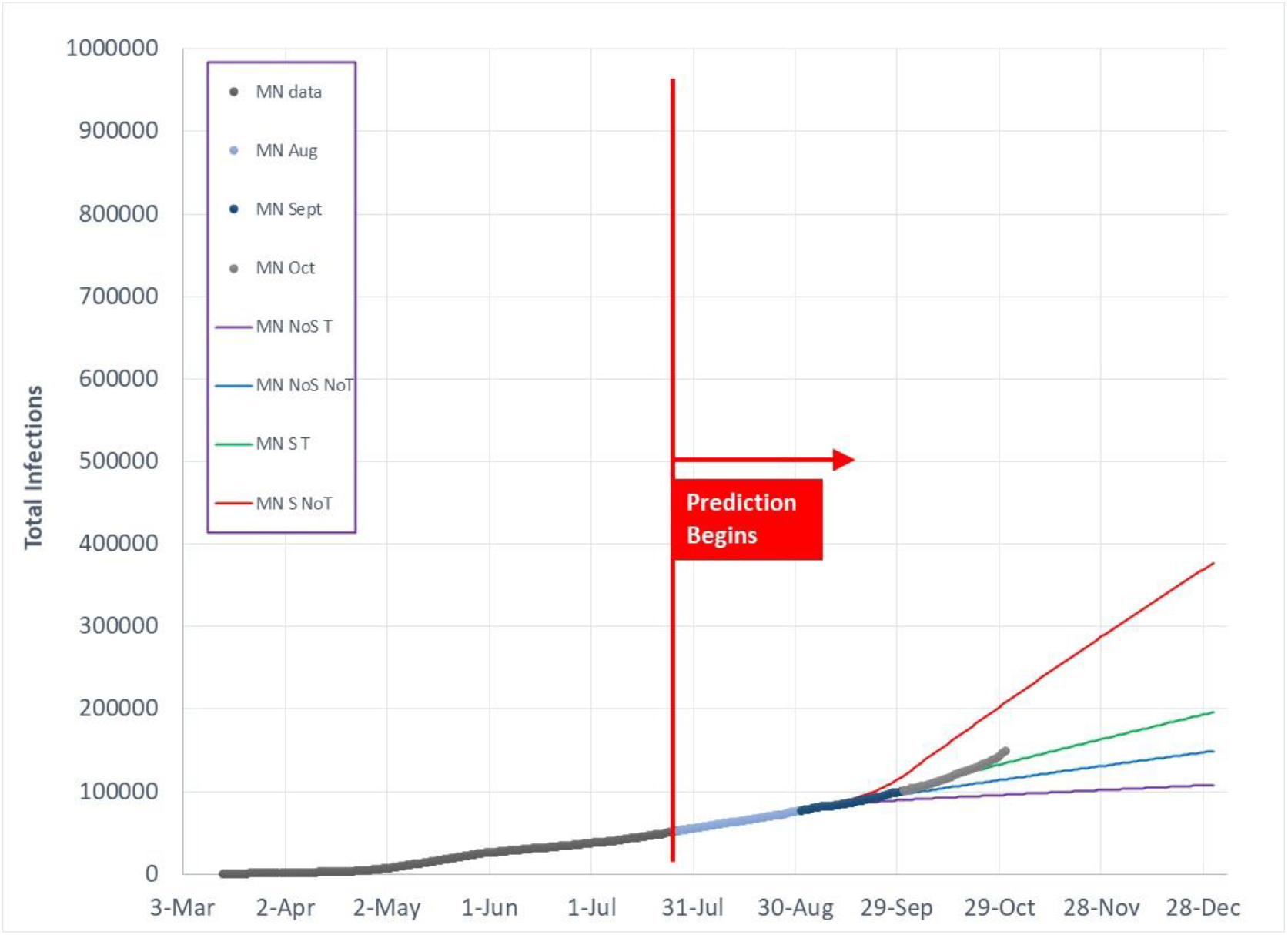
Total Covid-19 infection predictions as of July 27, 2020 for 4 model cases in comparison to actual data for MN.

**Figure 11.**
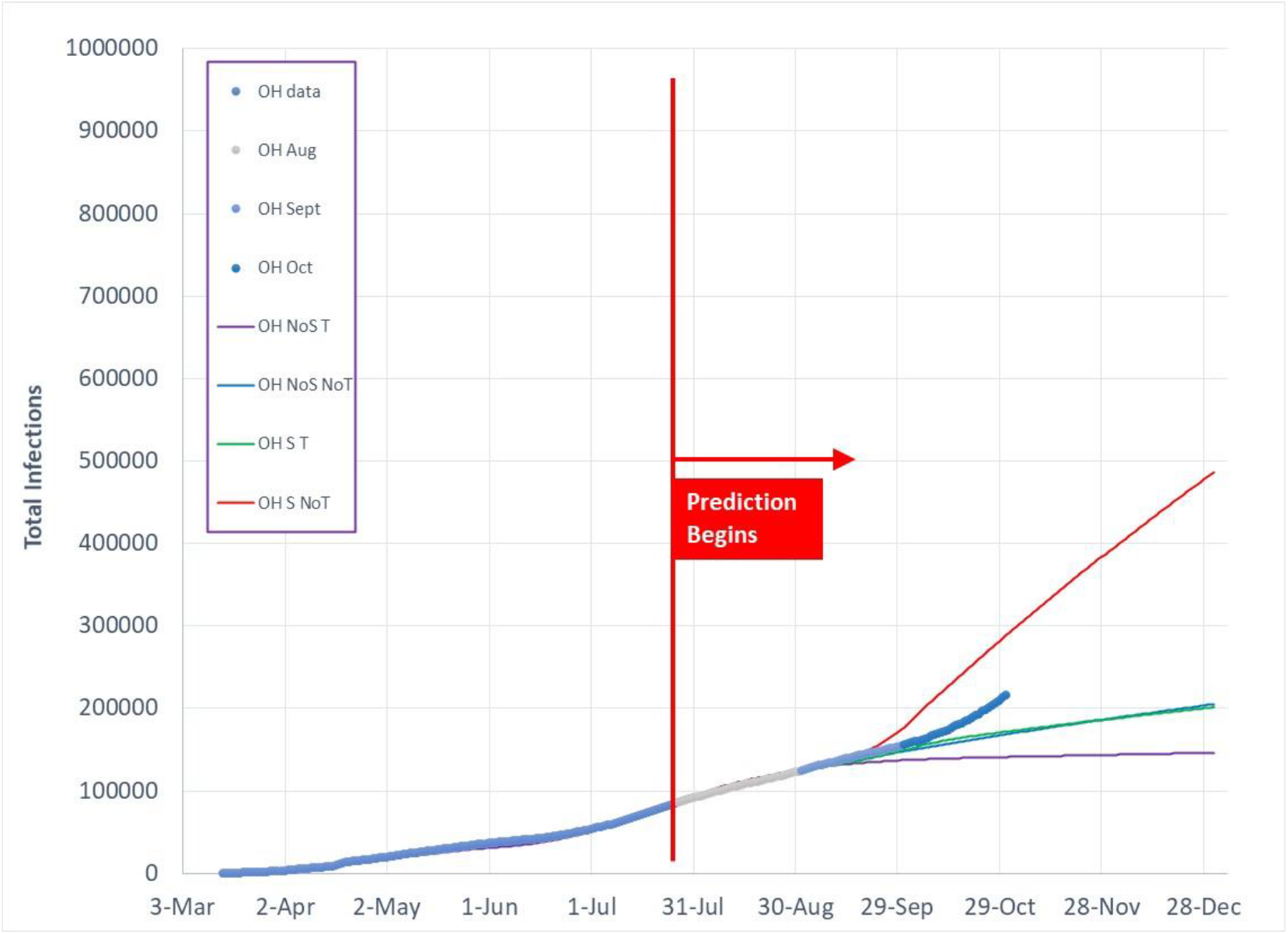
Total Covid-19 infection predictions as of July 27, 2020 for 4 model cases in comparison to actual data for OH.

**Figure 12.**
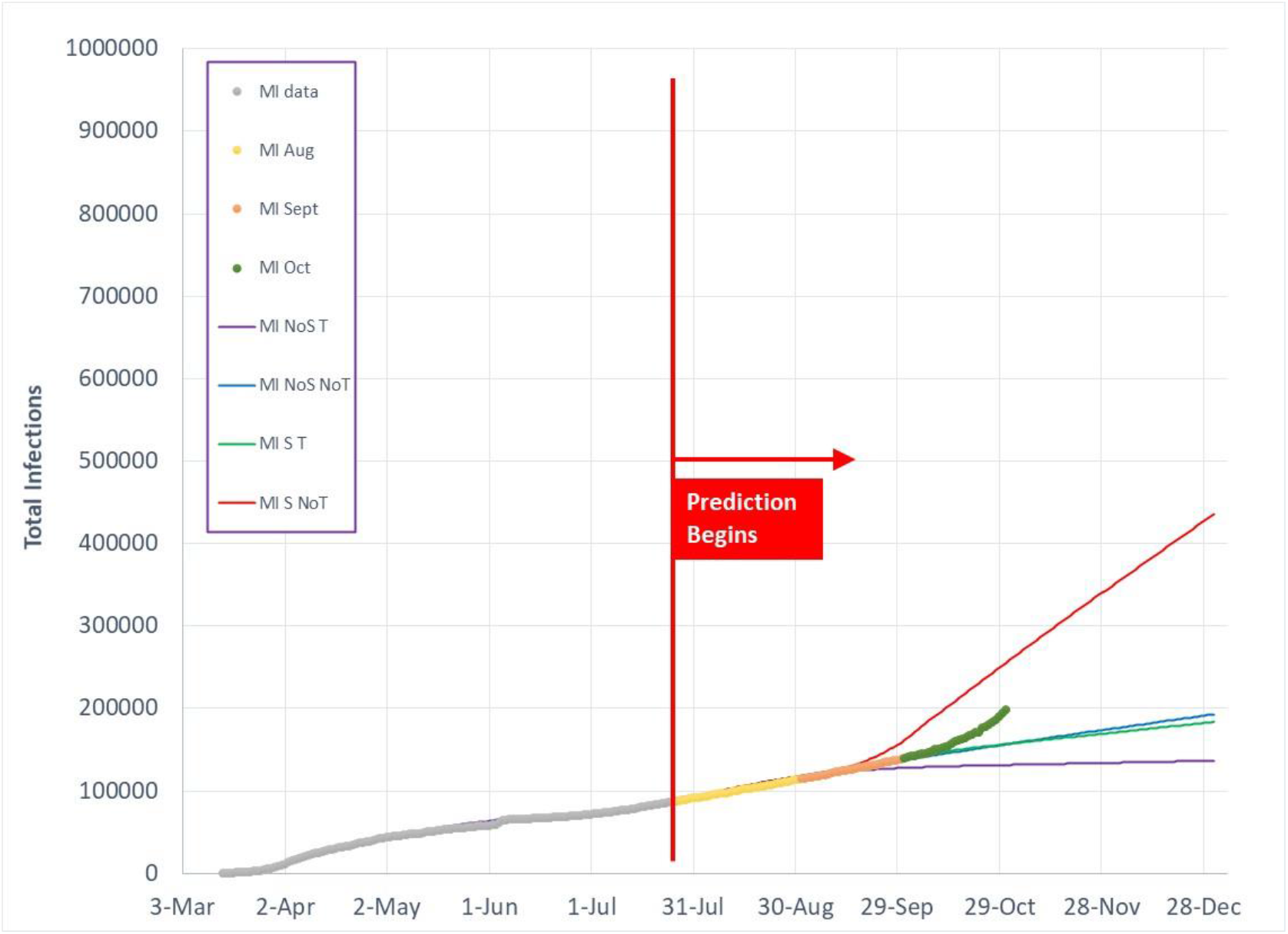
Total Covid-19 infection predictions as of July 27, 2020 for 4 model cases in comparison to actual data for MI.

Figures 13, 14, and 15 display FL, CA and WA infection trends. Florida (Figure 13) does not have a strong outdoor temperature impact as seen in more northerly states, which results in two paths for the four cases based on school openings. Through October, FL has followed the “no school” path, which is a continuation of Florida’s reaction to a strong summer surge. As a result of the summer surge (2,3), FL sustained an IP below the linear 2.72 boundary. As of the end of October, FL continued to have relatively high SDI, indicating a cautious movement toward increased social interactions. By the end of October, however, Florida’s infection path is curving upward and crossing into a range indicating increased social interaction and relaxation of local behavior (ie, reduced face mask usage, less distancing).

**Figure 13.**
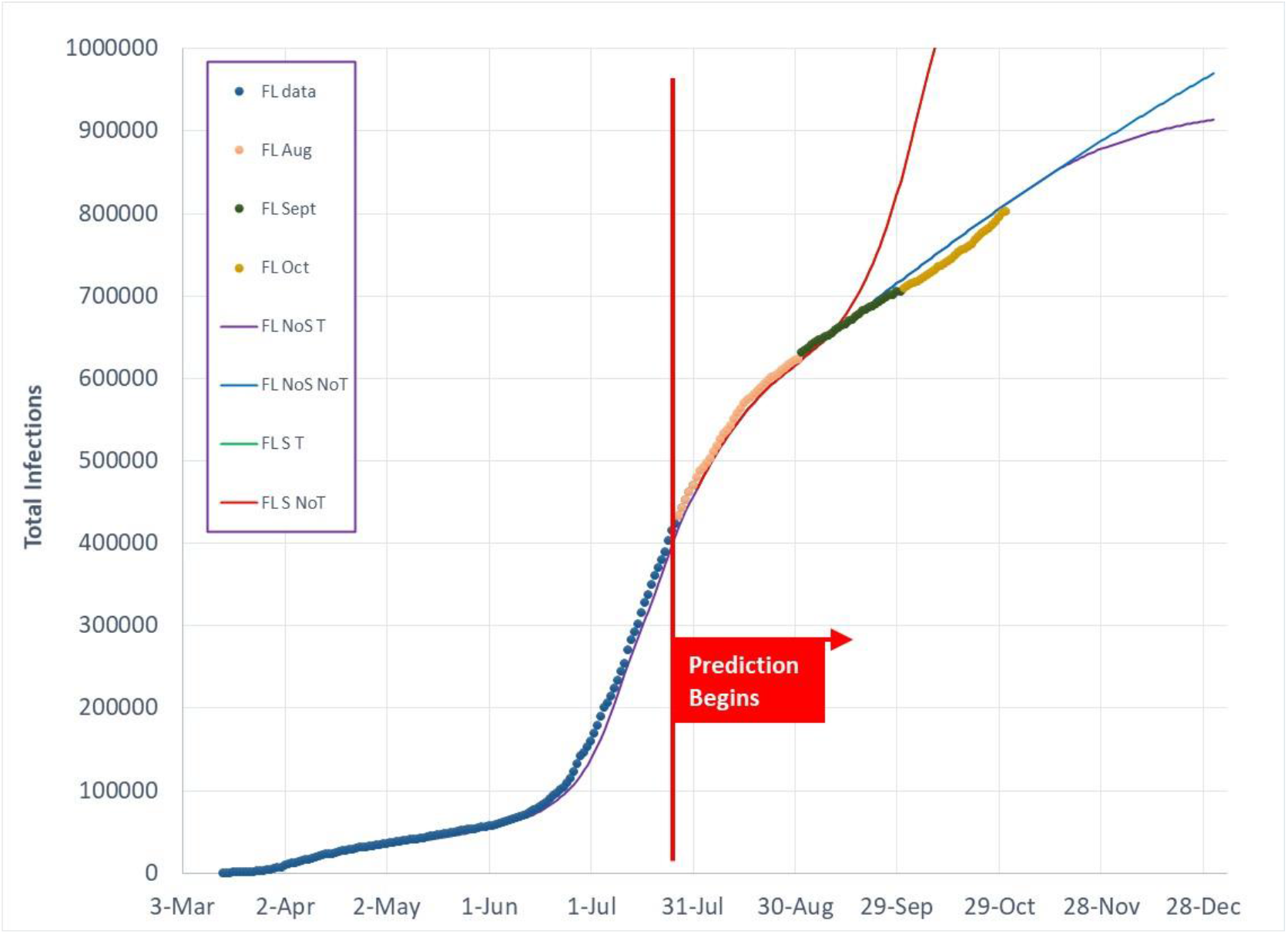
Total Covid-19 infection predictions as of July 27, 2020 for 4 model cases in comparison to actual data for FL.

**Figure 14.**
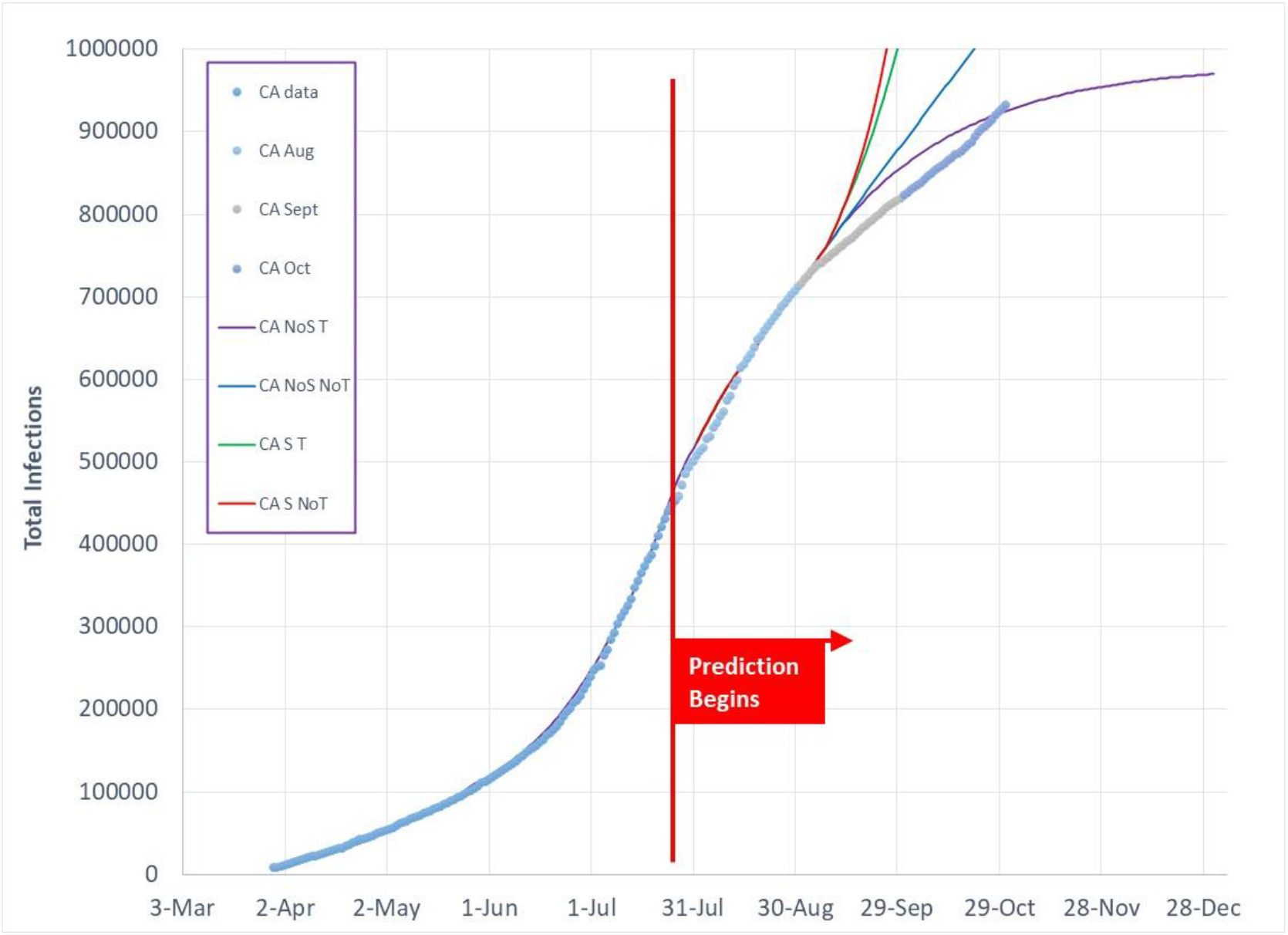
Total Covid-19 infection predictions as of July 27, 2020 for 4 model cases in comparison to actual data for CA.

**Figure 15.**
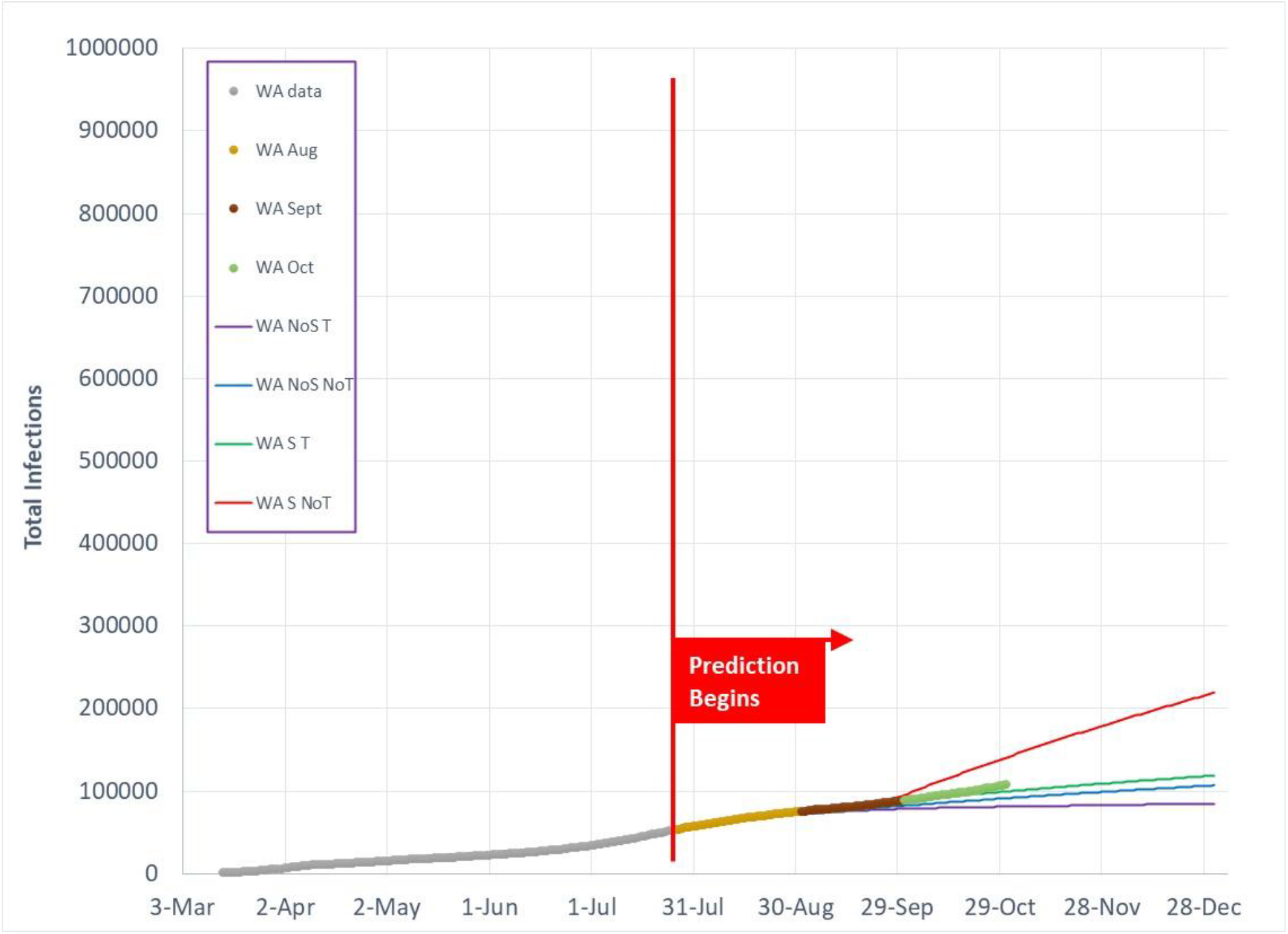
Total Covid-19 infection predictions as of July 27, 2020 for 4 model cases in comparison to actual data for WA.

California (Figure 14) has followed a similar trend as Florida. As a result of the summer surge, California reacted with strong infection control measures that continued through September. As seen in Figure 14, California’s infection growth was slightly below the four assumed model cases, however, by the end of October, California appeared to be relaxing its control measures and is now crossing into the range predicted for infection growth.

Washington (Figure 15) does not reach average summer temperatures above 70F (22C), although it is experiencing more frequent hot weather spells and air conditioning is increasing in homes and buildings. Because Washington stays in the beneficial outdoor temperature through the summer until the end of October, infection growth rates stayed relatively low. As Washington’s outdoor temperature drops below 50F (10C) at the end of October, infection growth rates are beginning to increase. Additional infection growth is expected in November as cold temperatures move people indoors coupled with increased social interactions.

## Summary

October has displayed predicted characteristics of accelerating infection growth due to a combination of increased social interactions coupled with increase disease transmission efficiency. Returning the US to the linear infection growth boundary (IP=2.72) separating accelerating infection growth from decaying infection transmission requires a combination of increased Social Distance Index by 15% and/or a decrease of disease transmission efficiency, G, by 27%. Additional SDI increase and G decrease must be sustained to reduce IP below 2.72, resulting in decay of new daily infections.

November will continue to be a challenging month of accelerating infection growth in the US due to incoherent, uncoordinated infection control strategies. The US will continue to oscillate across the linear infection growth boundary until a sustained infection transmission control effort is instituted.

## Data Availability

All data used for analyses are publicly available
1)Maryland Transportation Institute (2020). University of Maryland COVID-19 Impact Analysis Platform, https://data.covid.umd.edu, University of Maryland, College Park, USA
2) World Covid-19 infection data; Worldometers.info
3) World and US Covid-19 data; 91-divoc.com, Prof Wade Fagen-Ulmschneider, Computer Science Dept, University of Illinois

https://www.worldometers.info/coronavirus/

http://91-divoc.com/

https://data.covid.umd.edu

## Notes

### Competing Interest Statement

The authors have declared no competing interest.

### Funding Statement

No funding supports this work

### Author Declarations

All appropriate research reporting guidelines have been followed. I confirm no clinical trials were conducted. I confirm that no IRB/oversight body approval is required for this work.

